# Clinical and genomic profiling of early-onset bladder cancer identifies key alterations and therapeutic targets

**DOI:** 10.1101/2025.01.10.25320337

**Authors:** Christopher J. Magnani, Vincent D. D’Andrea, Guilherme Garcia Barros, Zhiyu Qian, John Ernandez, Kendrick Yim, Adam S. Kibel, Steven L. Chang, Matthew Mossanen, Mark A. Preston, Adam S. Feldman, Bernard H. Bochner, David B. Solit, Eugene J. Pietzak, Kent W. Mouw, Filipe L.F. Carvalho, Timothy N. Clinton

## Abstract

**Purpose:** Younger patients with bladder cancer typically have fewer environmental exposures or risk factors, suggesting that the molecular pathways implicated in early-onset disease differ from those in older patients. Dissecting the genomic profiles of early-onset tumors may inform targeted treatment strategies for young patients. We compared the frequency of somatic mutations in early-onset bladder cancer, defined as a diagnosis before the age of 55 years, which encompasses the youngest patients in about 1 in 10 diagnoses based on national registries.

**Material and methods:** We assessed two institutional cohorts: the Mass General Brigham Young Cystectomy Cohort (MGB-YCC, n=134 patients) from 2008-2023 and the Memorial Sloan-Kettering-IMPACT bladder cancer cohort (MSK-IMPACT, n=1271 including n=207 cystectomy and n=811 transurethral resection (TUR) specimens from 2014-2021. Clinical outcomes of MGB-YCC were also compared with published data from patients undergoing cystectomy in the National Surgical Quality Improvement Program (NSQIP, n=10,848) from 2008-2013.

**Results:** Both the MGB-YCC and MSK-IMPACT cohorts were predominantly male (>75%), and younger patients had lower rates of smoking history or muscle-invasive disease. MGB-YCC demonstrated more patients with continent diversions (51.5% vs 14.9%) than the older population in NSQIP, although complication rates were similar. Genomic characterization of MGB-YCC demonstrated enrichment in younger patients of TP53 (65% overall, 83% <50-year-old, p=0.045) and *KMT2D* mutations almost exclusively in the youngest patients (35% overall, 83% <45-years-old, p=0.008). MSK-IMPACT revealed a significant increase in *FGFR3* mutations in younger patients in both cystectomy (33% vs 13%, p=0.014) and TUR (45% vs 27%, p<0.001).

**Conclusions:** Our results indicate that early-onset bladder cancer is a distinct patient population that has disease driven by specific somatic mutations, some of which represent therapeutic targets. This suggests potential benefits of genomic tumor profiling in guiding personalized treatment.

## Introduction

Bladder cancer is the sixth most common malignancy in the United States, with an estimated 84,530 new cases and 17,870 deaths by 2026^1^. Despite advances in surgical management and systemic therapies, urothelial carcinoma (UC) remains a major cause of cancer-related morbidity and mortality. Environmental and lifestyle risk factors, including tobacco use and occupational carcinogen exposure to aromatic amines, account for a substantial proportion of disease burden^2-4^. Consistent with the cumulative nature of these exposures, the incidence of bladder cancer markedly increases with age. The median age at diagnosis is 73 years, and more than 90% of patients are diagnosed after age 55^1, 5^. However, early-onset bladder cancer represents a clinically important subset with unique biological and survivorship features. Previous studies have shown that younger patients are diagnosed more frequently with low-grade tumors and experience favorable long-term outcomes^6^. Nonetheless, when younger patients present with muscle-invasive or metastatic disease, tumors have a more aggressive behavior with increased rates of multifocal spread and a greater propensity for visceral metastases beyond the lymph nodes, including liver and brain involvement^7^. These results suggest that aggressive early-onset disease reflects distinct oncogenic mechanisms rather than earlier detection of urothelial carcinoma.

Younger bladder cancer patients have had less time to accumulate carcinogenic insults from smoking and environmental exposures, suggesting that tumorigenesis may rely on earlier acquisition of key driver somatic mutations or inherited germline alterations^2, 8^. Familial clustering of bladder cancer has been reported in some cases and appears independent of tobacco exposure, further supporting the contribution of germline or early life genomic events^9^. Moreover, a bladder cancer diagnosis at a young age carries disproportionate psychosocial and economic impacts, including fertility concerns, sexual dysfunction, and the need for decades of surveillance and survivorship care^10^. Progress in understanding early-onset bladder cancer has been limited in part by inconsistent definitions across studies, with widely variable age cutoffs that hinder cross-cohort comparisons and collaborative investigation^1,5^.

Given the reduced cumulative exposure to traditional carcinogens, we hypothesized that early-onset bladder cancer, particularly in patients presenting with aggressive disease, may be driven by distinct molecular pathways and potentially enriched for actionable genomic alterations. Over the past decade, large-scale genomic profiling studies have substantially advanced our understanding of the biology of urothelial carcinoma across disease states. Comprehensive sequencing efforts have identified recurrent alterations in chromatin remodeling, DNA damage response, receptor tyrosine kinase signaling, and cell-cycle regulation^11-14^. These insights have led to increasingly informed therapeutic development, including biomarker-driven approaches targeting DNA repair vulnerabilities^8, 15^. Clinical trials such as ATLANTIS and BAYOU have highlighted the growing relevance of molecular stratification in advanced urothelial carcinoma^16, 17^. However, despite these advances, most genomic studies have not focused on diagnostic age, and the molecular landscape of bladder cancer in younger patients remains incompletely characterized.

Therefore, in this study, we performed comprehensive clinical and genomic profiling of young patients with aggressive early-onset bladder cancer to nominate somatic mutations and therapeutic vulnerabilities present in this subset of bladder tumors. By dissecting early-onset bladder cancer biology, we aim to advance precision oncology strategies and stimulate collaborative efforts focused on this unique population with highly unmet clinical needs.

## Materials and Methods

### Study Design and Patient Population

We conducted a retrospective, multi-institutional cohort study (Massachusetts General Hospital, Brigham and Women’s Hospital, Memorial Sloan Kettering Cancer Institute) to characterize the clinical and genomic landscapes of early-onset bladder cancer. Early-onset disease was defined as a diagnosis before the age of 55 years, corresponding to approximately the youngest 10% of newly diagnosed bladder cancer patients based on national registry data^1, 5^. This cut-off was selected to provide a clinically meaningful and reproducible definition that enables comparisons across institutional cohorts.

### Mass General Brigham Young Cystectomy Cohort (MGB-YCC)

The primary institutional cohort consisted of patients treated within the Mass General Brigham system (Massachusetts General Hospital and Brigham and Women’s Hospital) who underwent radical cystectomy (RC) with curative intent between 2008 and 2023. Patients were included if they had a primary diagnosis of bladder cancer and were younger than 55 years of age at the time of diagnosis. A total of 182 patients aged <55 years who underwent RC were initially identified. Exclusion criteria were applied to ensure a homogeneous cohort of bladder cancer cases treated with curative intent. Specifically, 4 patients undergoing palliative cystectomy were excluded, as were 44 patients who underwent pelvic exenteration for non–bladder cancer primary malignancies. After exclusions, the final Mass General Brigham Young Cystectomy Cohort (MGB-YCC) included 134 patients. Clinical and pathological variables were collected through structured chart review and included demographic characteristics, tumor stage and histology, perioperative treatment, and oncologic outcomes.

A subset of patients within the MGB-YCC cohort underwent targeted next-generation sequencing of tumor tissue as part of routine clinical care. Sequencing was performed using the institutional OncoPanel platform, a clinically validated assay designed to detect actionable somatic alterations across cancer-associated genes^18^. Genomic findings from this subset (n=17) were analyzed descriptively to assess for potential enrichment of driver mutations in early-onset aggressive disease.

### MSK Integrated Mutation Profiling of Actionable Cancer Targets (MSK-IMPACT) Cohort

Given the small size of the study, we assessed an additional exploratory cohort from an independent external dataset to allow some comparisons of somatic genomic alterations. Genomic and clinical data were obtained through cBioPortal from the Memorial Sloan Kettering Integrated Mutation Profiling of Actionable Cancer Targets (MSK-IMPACT) cohort^11^. This dataset includes tumor-normal matched targeted sequencing with paired blood germline samples collected between 2014 and 2021. A total of 1,313 bladder cancer patients were available. After excluding 42 patients with missing diagnostic age information, the final analytic cohort included 1,271 patients of whom 212 (16.7%) were diagnosed before the age of 55 years. This cohort included 207 cystectomy and 811 transurethral resection (TUR) specimens, with 27 (13%) and 143 (18%) meeting criteria for early-onset bladder cancer, respectively. Somatic alterations and pathway-level genomic features were compared between early-onset and later-onset cases.

### National Surgical Quality Improvement Program

Outcomes from the MGB-YCC cohort were compared with published benchmark data from the National Surgical Quality Improvement Program (NSQIP) to contextualize the institutional results. This reference cohort included 10,848 patients undergoing radical cystectomy between 2008 and 2013, as previously reported^19^. Comparisons between MGB-YCC and NSQIP focused on perioperative and clinical outcomes in younger patients undergoing cystectomy.

### Statistical Analysis

Clinical and genomic comparisons were performed using R version 4.3.2 (R Foundation for Statistical Computing, Vienna, Austria). Categorical variables were compared using chi-square tests, while continuous variables were evaluated using Kruskal-Wallis tests. All statistical tests were two-sided, and a p-value <0.05 was considered statistically significant.

### Ethical Approval

This study was conducted in accordance with institutional and ethical standards for human subject research. This project was approved by the Mass General Brigham Institutional Review Board (IRB protocol 2021P002957). Patient data were retrospectively analyzed with appropriate safeguards for confidentiality.

## Results

### Patient Characteristics and Clinical Features of Early-Onset Bladder Cancer

Across both institutional and external cohorts, patients with early-onset bladder cancer (diagnosis <55 years) were predominantly male and frequently presented with a clinically aggressive disease. In the Mass General Brigham Young Cystectomy Cohort (MGB-YCC; n=134), 84% of patients were male, while in MSK-IMPACT 81% of cystectomies (n=167) and 70% with early-onset cases (n=19) were male (Tables 1–2, 4, S1– S2). The rates of muscle-invasive disease (_≥_T2) in early-onset cases undergoing cystectomy were comparable between cohorts, occurring in 57% and 63% of MGB-YCC and MSK-IMPACT patients, respectively. Smoking exposure was also common, reported in 55% and 59%, respectively (Tables 1–2, 4). Among young patients undergoing RC in the MGB-YCC cohort, multimodal treatment and contemporary surgical approaches were frequently used. Neoadjuvant chemotherapy was administered in 56% of cases, and urinary diversion patterns demonstrated a high utilization of continent reconstruction (51.5%) and robotic-assisted (9%) procedures (Table 1, Table S1).

**Table 1.** MGB Young Cystectomy Cohort (MGB-YCC) Patient Characteristics. Demographic features, treatment history and tumor characteristics of patients in MGB-YCC cohort. Early-onset patients stratified in subgroups younger than 45 years old, 45-49, and 50-54 years old. Chi-square test, p-value <0.05 was considered statistically significant.

| Age | Overall | < 45 | 45 - 49 | 50 - 54 | p |
| --- | --- | --- | --- | --- | --- |
| N (%) | 134 (100.0) | 31 (23.1) | 48 (35.8) | 55 (41.0) |  |
| Age, median [IQR] | 49 [45, 52] | 41 [37, 43] | 48 [46, 49] | 52 [51, 53] | <0.001 |
| Follow up - months, median [IQR] | 46 [16, 89] | 48 [25, 104] | 25 [12, 68] | 49 [26, 99] | 0.04 |
| BMI<sup>b</sup>, mean (SD) | 28.91 (5.99) | 29.29 (6.48) | 29.48 (6.11) | 28.18 (5.61) | 0.5 |
| Male (%) | 113 (84.3) | 25 (80.6) | 42 (87.5) | 46 (83.6) | 0.7 |
| Race = White (%) | 129 (96.3) | 28 (90.3) | 48 (100.0) | 53 (96.4) | 0.09 |
| Smoking (%) | 73 (54.5) | 15 (48.4) | 25 (52.1) | 33 (60.0) | 0.5 |
| Stage ≥T2, Muscle Invasive (%) | 76 (56.7) | 15 (48.4) | 34 (70.8) | 27 (49.1) | 0.048 |
| Ileal Conduit (%) | 65 (48.5) | 13 (41.9) | 22 (45.8) | 30 (54.5) | 0.5 |
| Robotic (%) | 12 (9.0) | 3 (9.7) | 6 (12.5) | 3 (5.5) | 0.5 |
| Neoadjuvant Chemo (%) | 75 (56.0) | 16 (51.6) | 26 (54.2) | 33 (60.0) | 0.7 |
| Length of stay - days, mean (SD) | 8.10 (6.04) | 7.68 (4.76) | 8.21 (5.78) | 8.25 (6.92) | 0.9 |
| Operative time - hours, median [IQR] | 8.1 [5.9, 9.6] | 8.2 [6.4, 10.0] | 8.4 [6.6, 9.7] | 6.9 [5.5, 9.0] | 0.1 |
| **Intravesical Treatment (%)** |  |  |  |  | 0.3 |
| None | 96 (71.6) | 22 (71.0) | 34 (70.8) | 40 (72.7) |  |
| BCG<sup>c</sup> | 23 (17.2) | 8 (25.8) | 9 (18.8) | 6 (10.9) |  |
| Chemo | 8 (6.0) | 0 (0.0) | 2 (4.2) | 6 (10.9) |  |
| Both | 7 (5.2) | 1 (3.2) | 3 (6.2) | 3 (5.5) |  |
| **Systemic Treatment (%)** |  |  |  |  | 0.8 |
| None | 49 (36.6) | 13 (41.9) | 17 (35.4) | 19 (34.5) |  |
| Chemo | 73 (54.5) | 15 (48.4) | 26 (54.2) | 32 (58.2) |  |
| Immunotherapy | 1 (0.7) | 0 (0.0) | 0 (0.0) | 1 (1.8) |  |
| Both | 11 (8.2) | 3 (9.7) | 5 (10.4) | 3 (5.5) |  |
| **Pathology, T Stage (%)** |  |  |  |  | 0.6 |
| T0 | 27 (20.1) | 8 (25.8) | 6 (12.5) | 13 (23.6) |  |
| Ta | 8 (6.0) | 2 (6.5) | 3 (6.2) | 3 (5.5) |  |
| Tis | 8 (6.0) | 1 (3.2) | 2 (4.2) | 5 (9.1) |  |
| T1 | 15 (11.2) | 5 (16.1) | 3 (6.2) | 7 (12.7) |  |
| T2 | 31 (23.1) | 4 (12.9) | 15 (31.2) | 12 (21.8) |  |
| T3 | 33 (24.6) | 9 (29.0) | 14 (29.2) | 10 (18.2) |  |
| T4 | 12 (9.0) | 2 (6.5) | 5 (10.4) | 5 (9.1) |  |

**Table 2.**
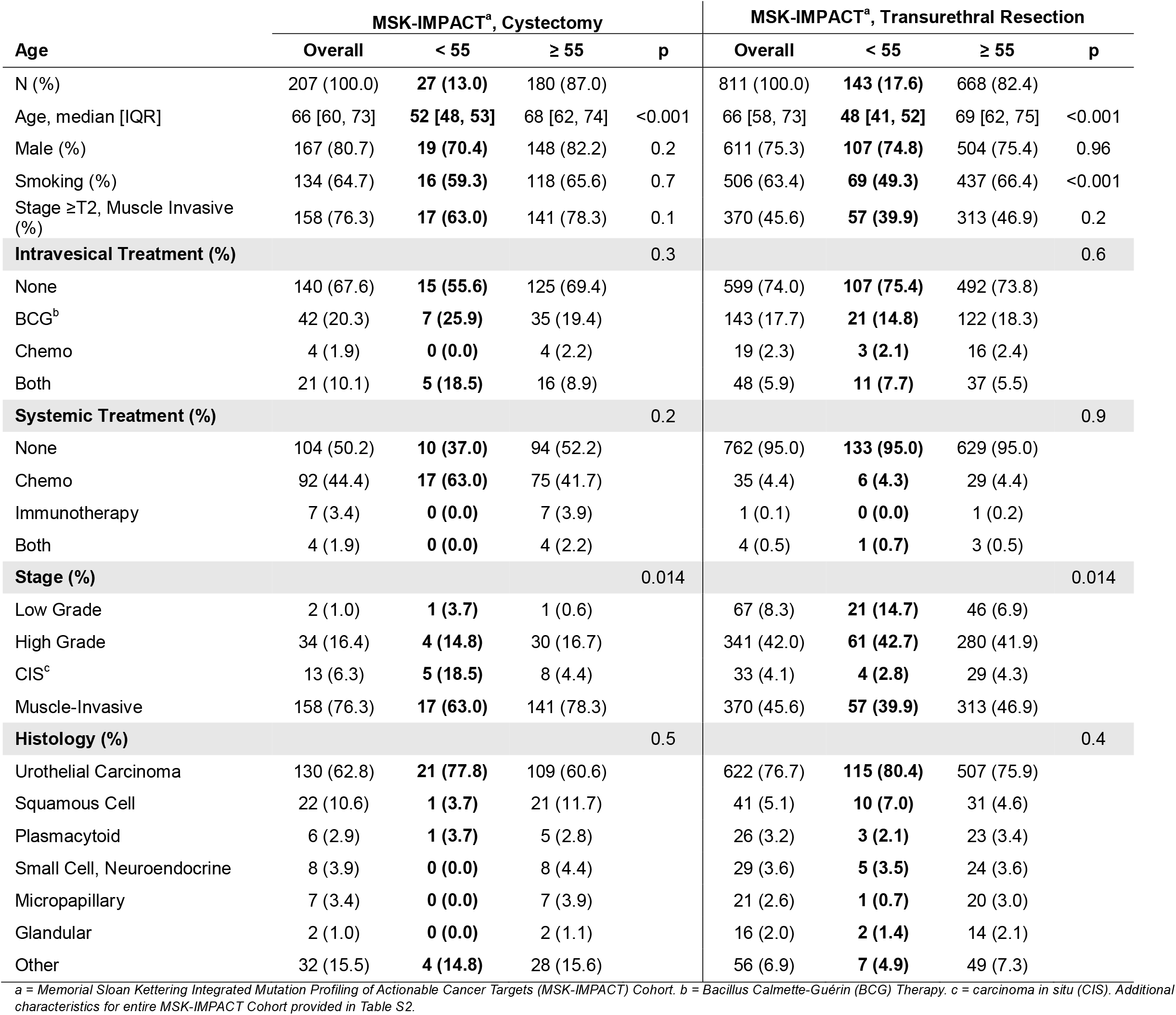
MSK-IMPACT Patient Characteristics. Demographic features, treatment history and tumor characteristics of patients in MSK-IMPACT cohort stratified by cystectomy and transurethral resection specimens. Comparisons made between patients with early-onset bladder cancer (age <55 years old) and older patients (age _≥_ 55). Chi-square test, p-value <0.05 was considered statistically significant.

**Table 3:**
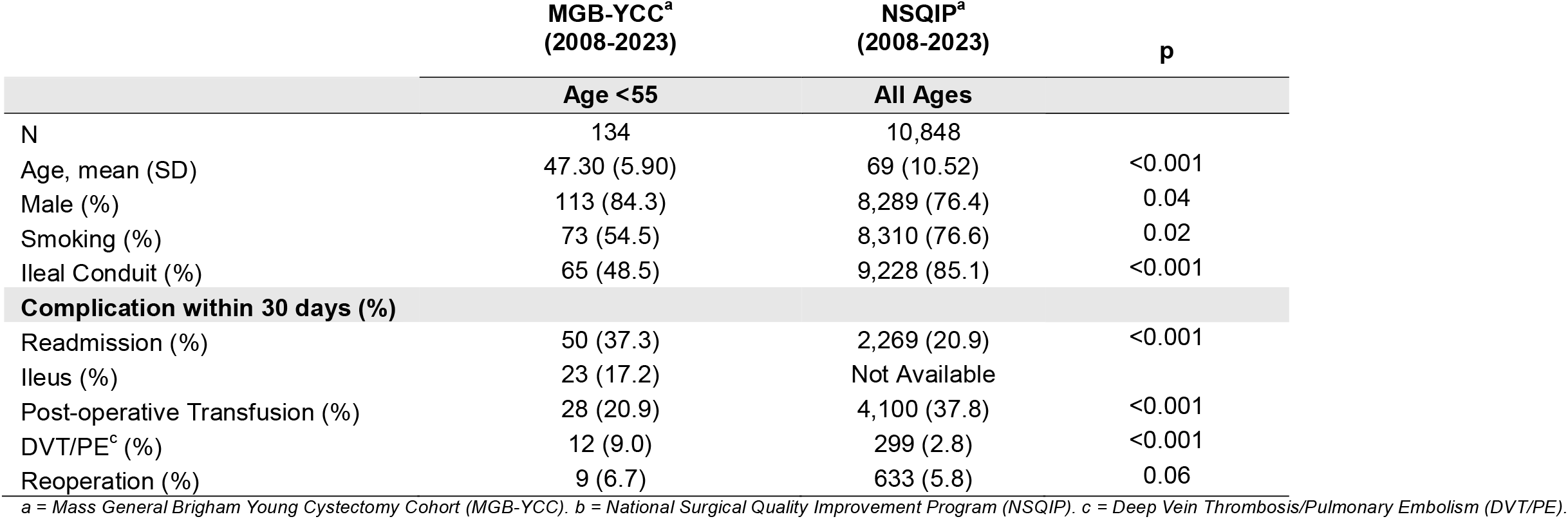
Post-Operative Complications. Comparison of complication rates after radical cystectomy in MGB-YCC and NSQIP. Fisher exact test, p-value <0.05 was considered statistically significant.

**Table 4:** Assessing Genomic Characteristics of Patients with Early-Onset Bladder Cancer. Comparison of characteristics and genomic mutation profiles of Mass General Brigham Young Cystectomy Cohort (MGB-YCC) and Memorial Sloan Kettering Integrated Mutation Profiling of Actionable Cancer Targets (MSK-IMPACT) cohort. MSK-IMPACT data presented for cystectomy and transurethral resection specimens in parallel. Early-onset bladder cancer defined as diagnosis before age 55. Additional somatic mutation characteristics provided in Tables S1-3 and Figure S1.

| Age | MGB-YCC<sup>b</sup> | MSK-IMPACT<sup>c</sup>, Cystectomy |  |  |  | MSK-IMPACT<sup>c</sup>, Transurethral Resection |  |  |  |
| --- | --- | --- | --- | --- | --- | --- | --- | --- | --- |
|  | Overall | Overall | < 55 | ≥ 55 | p | Overall | < 55 | ≥ 55 | p |
| N (%) | **134 (100.0)** | 207 (100.0) | **27 (13.0)** | 180 (87.0) |  | 811 (100.0) | **143 (17.6)** | 668 (82.4) |  |
| Age, median [IQR] | **49 [45, 52]** | 66 [60, 73] | **52 [48, 53]** | 68 [62, 74] | <0.001 | 66 [58, 73] | **48 [41, 52]** | 69 [62, 75] | <0.001 |
| Male (%) | **113 (84.3)** | 167 (80.7) | **19 (70.4)** | 148 (82.2) | 0.2 | 611 (75.3) | **107 (74.8)** | 504 (75.4) | >0.9 |
| Smoking (%) | **73 (54.5)** | 134 (64.7) | **16 (59.3)** | 118 (65.6) | 0.7 | 506 (63.4) | **69 (49.3)** | 437 (66.4) | <0.001 |
| Stage ≥T2 (%) | **76 (56.7)** | 158 (76.3) | **17 (63.0)** | 141 (78.3) | 0.1 | 370 (45.6) | **57 (39.9)** | 313 (46.9) | 0.2 |
| **Highlighted Mutations** |  |  |  |  |  |  |  |  |  |
| TP53 | **11 (64.7)** | 110 (53.1) | **12 (44.4)** | 98 (54.4) | 0.4 | 367 (45.3) | **53 (37.1)** | 314 (47.0) | 0.038 |
| KMT2D | **6 (35.3)** | 37 (17.9) | **2 (7.4)** | 35 (19.4) | 0.2 | 157 (19.4) | **26 (18.2)** | 131 (19.6) | 0.8 |
| ARID1A | **4 (23.5)** | 34 (16.4) | **4 (14.8)** | 30 (16.7) | >0.9 | 198 (24.4) | **27 (18.9)** | 171 (25.6) | 0.1 |
| RB1 | **4 (23.5)** | 37 (17.9) | **3 (11.1)** | 34 (18.9) | 0.5 | 161 (19.9) | **18 (12.6)** | 143 (21.4) | 0.022 |
| TERT | **4 (23.5)** | 131 (63.3) | **15 (55.6)** | 116 (64.4) | 0.5 | 620 (76.4) | **97 (67.8)** | 523 (78.3) | 0.010 |
| KDM6A | **4 (23.5)** | 53 (25.6) | **7 (25.9)** | 46 (25.6) | >0.9 | 217 (26.8) | **36 (25.2)** | 181 (27.1) | 0.7 |
| PIK3CA | **3 (17.6)** | 27 (13.0) | **5 (18.5)** | 22 (12.2) | 0.5 | 165 (20.3) | **31 (21.7)** | 134 (20.1) | 0.7 |
| CDKN1A | **2 (11.8)** | 22 (10.6) | **3 (11.1)** | 19 (10.6) | >0.9 | 104 (12.8) | **20 (14.0)** | 84 (12.6) | 0.7 |
| FGFR3 | **1 (5.9)** | 32 (15.5) | **9 (33.3)** | 23 (12.8) | 0.014 | 243 (30.0) | **64 (44.8)** | 179 (26.8) | <0.001 |
| CDKN2A | **1 (5.9)** | 42 (20.3) | **4 (14.8)** | 38 (21.1) | 0.6 | 178 (21.9) | **25 (17.5)** | 153 (22.9) | 0.2 |
| STAG2 | **1 (5.9)** | 22 (10.6) | **2 (7.4)** | 20 (11.1) | 0.8 | 89 (11.0) | **18 (12.6)** | 71 (10.6) | 0.6 |
| CDKN2B | **0 (0.0)** | 25 (12.1) | **3 (11.1)** | 22 (12.2) | >0.9 | 146 (18.0) | **23 (16.1)** | 123 (18.4) | 0.6 |
| CCND1 | **0 (0.0)** | 19 (9.2) | **3 (11.1)** | 16 (8.9) | >0.9 | 89 (11.0) | **17 (11.9)** | 72 (10.8) | 0.8 |
| ERBB2 | **0 (0.0)** | 26 (12.6) | **5 (18.5)** | 21 (11.7) | 0.5 | 114 (14.1) | **12 (8.4)** | 102 (15.3) | 0.044 |
| EP300 | **0 (0.0)** | 14 (6.8) | **2 (7.4)** | 12 (6.7) | >0.9 | 62 (7.6) | **14 (9.8)** | 48 (7.2) | 0.4 |

*a = Early-onset bladder cancer defined as diagnosis before age 55, based on youngest 10% of patients with new diagnoses in national registry data<sup>18</sup>. b = Mass General Brigham Young Cystectomy Cohort (MGB-YCC). c = Memorial Sloan Kettering Integrated Mutation Profiling of Actionable Cancer Targets (MSK-IMPACT) Cohort.*

### Clinical Comparison of Early-Onset Versus Older-Onset Disease in MSK-IMPACT

Clinical and pathological features of patients with early-onset bladder cancer patients (diagnostic age <55 years) were compared with those of patients with later-onset disease (diagnostic age _≥_55) to assess age-associated differences and stratified between cystectomy and TUR specimens. Early-onset patients demonstrated a similar sex distribution to that of later-onset patients (70%-82% male). However, smoking prevalence was significantly lower among TUR cases from younger individuals (49% vs 66%, p<0.001) though similar trends were not significant among cystectomies (59% vs 66%, p=0.7), supporting the reduced cumulative carcinogen exposure in this population (Tables 2, S2). Younger patients demonstrated less muscle-invasive disease (63% vs 78%, p=0.01) in cystectomy specimens and a greater proportion of low-grade tumors (15% vs 7%, p=0.01) in TUR specimens, consistent with prior studies suggesting heterogeneity in clinical behavior among early-onset cases (Tables 2, S2). Variant histology was less common in early-onset disease in both cystectomy and TUR specimens, though this was not significant.

### Perioperative Outcomes Compared with National Benchmarks

To contextualize outcomes in young cystectomy patients, we compared perioperative results from MGB-YCC with national benchmark data from NSQIP (n=10,848 radical cystectomy patients of all ages)^19^. The NSQIP cohort demonstrated a predominantly male population (76.4%), although smoking prevalence was substantially higher (76.6%) than that in the younger institutional cohort (Table 3). Additionally, continent diversion was far less common nationally (14.9%), consistent with older age distributions and differing practice patterns (Table 3). Thirty-day postoperative complication rates were generally comparable between cohorts. However, MGB-YCC demonstrated slightly higher rates of reported readmission (37% vs 21%), deep venous thrombosis/pulmonary embolism (DVT/PE, 9% vs 3%), and reoperation (7% vs 6%) (Table 3). These differences reflect the more granular capture of postoperative events in the institutional cohort compared with registry-based reporting rather than true excess morbidity among younger patients.

### Genomic Landscape of Early-Onset Bladder Cancer

Genomic profiling of early-onset bladder cancer revealed distinct somatic alterations across cohorts (Table 4). In the MGB-YCC sequencing subset, somatic mutations in *TP53* (65%) and *KMT2D* (35%) were significantly enriched in the youngest patients (83% age<50 p=0.045 and 83% age<45 p=0.008, respectively; Table S1). These findings suggest that alterations in the tumor suppressor and chromatin remodeling pathways may play a prominent role in aggressive early onset disease requiring cystectomy. Age-associated differences in recurrent driver alterations were also observed in the MSK-IMPACT cohort. Early-onset patients demonstrated a significant enrichment of somatic *FGFR3* mutations in both cystectomy (33% vs 13%, p=0.014) and TUR cases (45% vs 27%, p<0.001), highlighting a potentially targetable oncogenic pathway in early-onset disease (Table 4, Figure 1). Conversely, early-onset tumors exhibited significantly lower mutation frequencies in several canonical urothelial carcinoma drivers, including *TERT* (RC 56% vs 64%, TUR 68% vs 78%), *TP53* (RC 44% vs 54%, TUR 37% vs 47%), and *RB1* (RC 11% vs 19%, TUR 13% vs 21%) (Table 4, Figure 1). These genomic relationships illustrate the MGB-YCC cohort’s enrichment of *KMT2D* somatic mutations in early-onset bladder cancer patients and the increased prevalence of *FGFR3* somatic mutations in MSK-IMPACT. Collectively, these findings support the existence of distinct molecular features in early-onset bladder cancer and suggest potentially actionable pathways that warrant further investigation.

**Figure 1:**
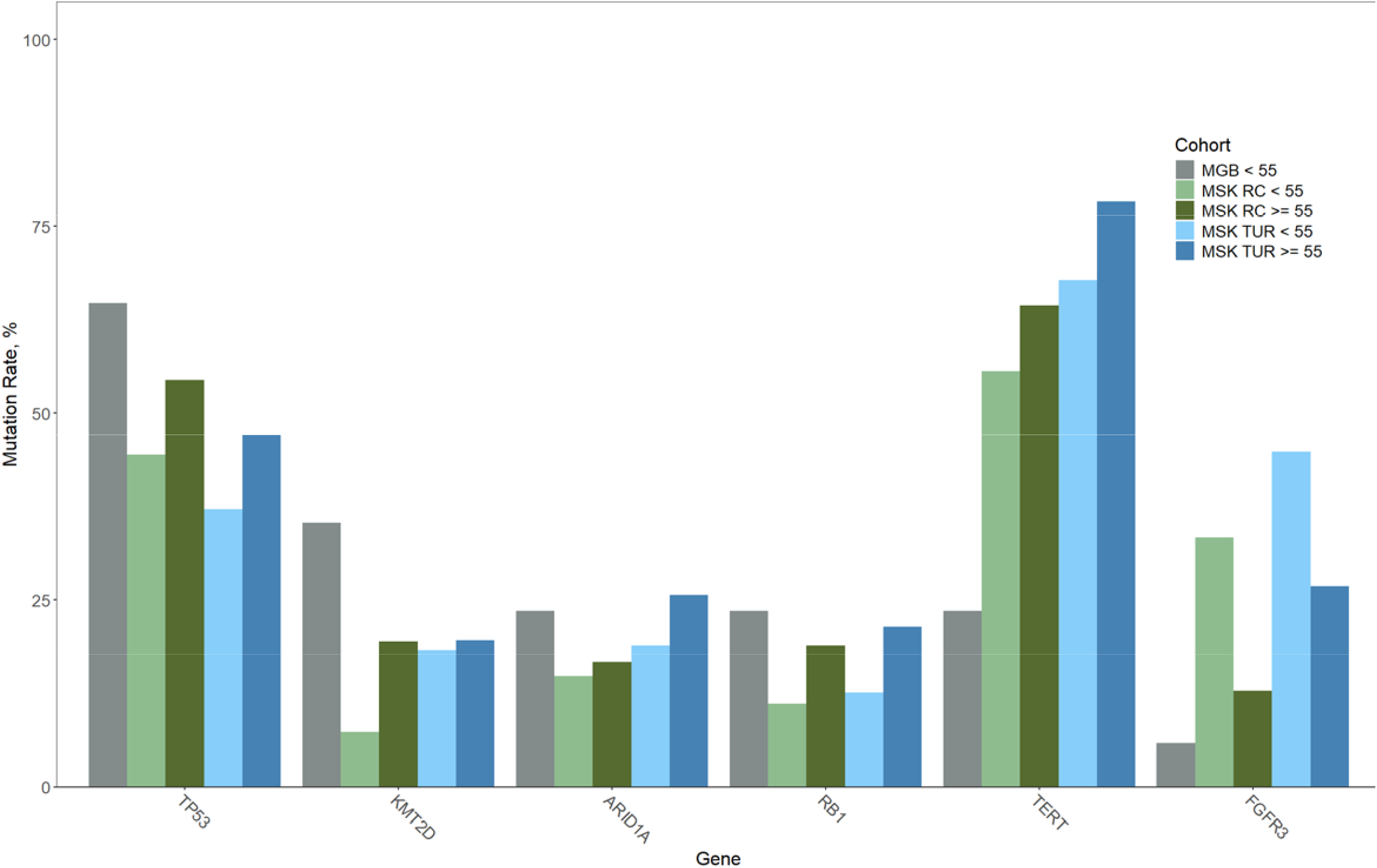
Somatic Mutation Rates: Bar chart with somatic genomic mutation rates of Mass General Brigham Young Cystectomy Cohort (MGB-YCC) and Memorial Sloan Kettering Integrated Mutation Profiling of Actionable Cancer Targets (MSK-IMPACT) cohort, which demonstrate a significant age-dependent distributions, also available in Table 4. Notably, KMT2D is enriched in MGB-YCC while MSK-IMPACT demonstrates FGFR3 enrichment in both cystectomy and transurethral resection specimens from younger patients.

## Discussion

Early-onset bladder cancer represents a clinically important subset of urothelial carcinoma. Here we defined early-onset as a bladder cancer diagnosis before the age of 55, corresponding to the youngest 10% of patients nationally^1, 5^. This population faces unique survivorship and psychosocial challenges, including prolonged treatment burden, fertility and sexual health issues, and decades of surveillance^10^. At the same time, emerging evidence suggests that bladder cancers diagnosed in younger individuals might be biologically distinct from tumors diagnosed later in life, particularly among young patients who present with aggressive disease^2, 7, 8^. However, the clinical and molecular drivers of aggressive early-onset bladder cancer remain incompletely characterized, limiting the development of age-informed precision oncology strategies.

In this study, we provide a comprehensive clinical and genomic assessment of early-onset bladder cancer in a multi-institutional exploratory analysis using two complementary cohorts. Our results showed that young patients with aggressive disease frequently underwent radical cystectomy with contemporary multimodal management, including high rates of neoadjuvant chemotherapy and substantial utilization of continent urinary diversion consistent with long-term quality of life goals. Importantly, overall perioperative complication rates in the MGB-YCC cohort were comparable to national NSQIP benchmarks, supporting the feasibility of aggressive surgical management in younger patients with curative intent. Although postoperative events such as readmission and venous thromboembolism were reported at higher rates in the MGB-YCC cohort, this potentially reflects a more complete capture of complications through a detailed chart review compared with registry-based reporting.

Notably, early-onset patients demonstrated lower smoking prevalence compared with later-onset cases in the MSK-IMPACT cohort, consistent with reduced cumulative exposure to established carcinogenic risk factors. These results support the hypothesis that aggressive bladder cancer arising at a young age may be less driven by prolonged environmental exposure and more dependent on earlier acquisition of key oncogenic alterations or inherited susceptibility. Familial clustering independent of smoking has been described in prior studies^9^, further reinforcing the need to investigate molecular pathways unique to young patients with bladder cancer.

Our study highlights the clinical heterogeneity of early-onset disease. Previous studies have shown that younger patients often present with more favorable low-grade tumors^6^, but both MGB-YCC and MSK-IMPACT cohorts represent patient populations with more aggressive tumor biology; the MGB-YCC cohort was defined by patients who underwent radical cystectomy, while MSK-IMPACT reflects both cystectomy and TUR patients with high-risk tumors referred to a tertiary academic center for specialized care. These observations reinforce that young patients with aggressive bladder cancer represent a subgroup of patients with an unmet need for biological discovery and therapeutic innovation.

A key contribution of this study is the identification of actionable genomic alterations enriched in early-onset bladder cancer. In the MGB-YCC cohort, we observed an increased frequency of *KMT2D* somatic mutations, implicating chromatin-remodeling pathways in aggressive early-onset disease. KMT2D encodes a histone methyltransferase with emerging relevance across tumor types, and epigenetic-targeted strategies are currently under investigation^20^. The MSK-IMPACT cohort analysis demonstrated a significant enrichment of *FGFR3* mutations in younger patients in both TUR and cystectomy patients. FGFR3 mutations are clinically actionable in urothelial carcinoma through the FGFR inhibitor erdafitinib, which is already FDA-approved for metastatic disease^21^. Together, these findings suggest that early-onset bladder cancer harbors distinct genomic alterations, and that tumor sequencing in young patients could have direct therapeutic relevance because some somatic mutations present in these tumors can be therapeutically targeted.

More broadly, ongoing collaborative consensus efforts are being made to provide a clear definition of early-onset bladder cancer using reproducible national thresholds and frameworks. Because this population represents a small proportion of patients with bladder cancer, meaningful advances in biologic understanding and clinical trial development will require coordinated efforts across institutions to combine genomic data, harmonize clinical annotation, and identify recurrent targetable pathways.

### Conclusions and Limitations

This study is one of the largest clinicogenomic analyses of early-onset bladder cancer and provides evidence of unique genomic alterations that can affect treatment selection. However, this study had several limitations. Both cohorts were derived from academic referral centers, which may enrich for aggressive or complex cases and limit generalizability to community-treated early-onset disease. In addition, sequencing within the MGB-YCC cohort was available only for a subset of patients, and differences in specimen type and clinical selection between cystectomy-treated and TUR-based cohorts may have influenced the observed genomic patterns. Furthermore, this work is only an exploratory analysis demonstrating genomic associations that would need to be rigorously tested at the molecular level to assess possible causal relationships. Despite these limitations, this represents one of the largest analyses to date that integrates clinical outcomes with genomic profiling in early-onset bladder cancer.

Our findings support the concept that early-onset disease, particularly in young patients with aggressive tumors, may exhibit distinct molecular features, including enrichment of actionable alterations such as FGFR3 and chromatin remodeling pathway involvement. Future multi-institutional studies with expanded sequencing and prospective clinical annotation will be essential to refine the genomic landscape of early-onset bladder cancer, validate therapeutic vulnerabilities, and ultimately enable personalized treatment strategies and dedicated clinical trials for this underserved population. It is our hope that exploratory analyses like ours will encourage greater interest in studying this population and stimulate collaborative opportunities to carry this work forward to the larger patient cohorts needed to strengthen understanding of molecular drivers in early-onset disease.

## Data Availability

Data are archived in an institutional database, these are not publicly available. Aggregate data are provided in the manuscript and statistical analysis and code can be discussed upon request.

## Abbreviations

BCG: Bacillus Calmette-Guérin Therapy
BMI: Body Mass Index
FDA: Food and Drug Administration
MGB-YCC: Mass General Brigham Young Cystectomy Cohort
MSK-IMPACT: Memorial Sloan Kettering Integrated Mutation Profiling of Actionable Cancer Targets
NCI: National Cancer Institute
NSQIP: National Surgical Quality Improvement Program
RC: Radical Cystectomy
TUR: Transurethral Resection
UC: Urothelial Carcinoma

## Legends

**Table S1: Detailed MGB-YCC clinical and tumor characteristics** . Detailed clinical characteristics, tumor features, and mutation profiles of early-onset bladder cancer cohort (age <55 years old). Chi-square test, p-value <0.05 was considered statistically significant.

**Table S2: Detailed MSK-IMPACT Patient Characteristics**. Detailed clinical characteristics, tumor features, and mutation profiles of early-onset bladder cancer cohort (age <55 years old). Chi-square test, p-value <0.05 was considered statistically significant. Full MSK-IMPACT cohort presented, see Table 2 for stratified data for cystectomy and transurethral resection specimens.

**Table S3: Variant Classifications for Most Frequent Mutations by Cohort**. Detailed counts and variant classifications for somatic mutation calls by cohort.

**Figure S1: Mutational landscape in the MGB-YCC cohort**. Comutation plot (left) presents somatic mutations profiled by MGB targeted sequencing panel (rows) in early-onset patients in the MGB-YCC cohort (columns). Variant classification (top right) demonstrates predominantly functional inactivating mutations, and provides color legend for mutation types across depicted panels (note black corresponds to multi-hit mutations), and is depicted individually for the 10 most mutated genes (bottom right). Detailed counts and variant classifications for mutation calls provided in Table S3.

**Figure S2: Mutational landscape in MSK-IMPACT cystectomy samples**. Stratified by age to distinguish early-onset (age <55 years old, upper set of panels) and late-onset (age _≥_55 years old, lower set of panels) patients. Comutation plot (left in each set) presents somatic mutations profiled by MSK-IMPACT targeted sequencing (rows) in cystectomy samples (columns). Variant classification (top right in each set) demonstrates predominantly functional inactivating mutations, and provides color legend for mutation types across depicted panels (note black corresponds to multi-hit mutations), and is depicted individually for the 10 most mutated genes (bottom right in each set). Detailed counts and variant classifications for mutation calls provided in Table S3.

**Figure S3: Mutational landscape in MSK-IMPACT transurethral resection samples**. Stratified by age to distinguish early-onset (age <55 years old, upper set of panels) and late-onset (age _≥_55 years old, lower set of panels) patients. Comutation plot (left in each set) presents somatic mutations profiled by MSK-IMPACT targeted sequencing (rows) in transurethral resection samples (columns). Variant classification (top right in each set) demonstrates predominantly functional inactivating mutations, and provides color legend for mutation types across depicted panels (note black corresponds to multi-hit mutations), and is depicted individually for the 10 most mutated genes (bottom right in each set). Detailed counts and variant classifications for mutation calls provided in Table S3.

## Notes

### Competing Interest Statement

The authors have declared no competing interest.

### Author Declarations

IRB of Mass General Brigham gave ethical approval for this work (IRB 2021P002957).

### Summary of Updates

Updated per recommendations during ongoing peer review process for eventual journal publication. The MSK-IMPACT cohort has now been separated to distinguish patient samples derived from cystectomy vs transurethral resection samples for more apt comparisons. Additionally, more detailed information is provided in the supplemental figures/tables for the gene mutation/variant classifications.

