## Supplementary material for "Clinical and genomic profiling of early-onset bladder cancer identifies key alterations and therapeutic targets": Table S1

**Table S1: Detailed MGB-YCC^a^ clinical and tumor characteristics**

| **Age** | **Overall** | **< 45** | **45 - 49** | **50 - 54** | **p** |
| --- | --- | --- | --- | --- | --- |
| N (%) | 134 (100.0) | 31 (23.1) | 48 (35.8) | 55 (41.0) |  |
| Age, mean (SD) | 47.30 (5.90) | 38.94 (5.93) | 47.35 (1.47) | 51.96 (1.41) | <0.001 |
| Age, median [IQR] | 49 [45, 52] | 41 [37, 43] | 48 [46, 49] | 52 [50.5, 53] | <0.001 |
| Follow up - months, mean (SD) | 59.06 (50.85) | 68.40 (53.70) | 45.94 (46.91) | 65.24 (51.07) | 0.08 |
| Follow up - months, median [IQR] | 45.8 [16.4, 89.1] | 48.3 [25.2, 103.5] | 25.4 [11.6, 68.4] | 48.6 [25.7, 99.1] | 0.04 |
| BMI^b^, mean (SD) | 28.91 (5.99) | 29.29 (6.48) | 29.48 (6.11) | 28.18 (5.61) | 0.5 |
| BMI^b^, median [IQR] | 28.6 [24.8, 32.0] | 27.1 [24.9, 32.9] | 29.8 [25.0, 32.2] | 27.2 [24.1, 31.0] | 0.5 |
| Male (%) | 113 (84.3) | 25 (80.6) | 42 (87.5) | 46 (83.6) | 0.7 |
| Race = White (%) | 129 (96.3) | 28 (90.3) | 48 (100.0) | 53 (96.4) | 0.09 |
| Smoking (%) | 73 (54.5) | 15 (48.4) | 25 (52.1) | 33 (60.0) | 0.5 |
| Stage ≥T2, Muscle Invasive (%) | 76 (56.7) | 15 (48.4) | 34 (70.8) | 27 (49.1) | 0.048 |
| Ileal Conduit (%) | 65 (48.5) | 13 (41.9) | 22 (45.8) | 30 (54.5) | 0.5 |
| Robotic (%) | 12 (9.0) | 3 (9.7) | 6 (12.5) | 3 (5.5) | 0.5 |
| Neoadjuvant Chemo (%) | 75 (56.0) | 16 (51.6) | 26 (54.2) | 33 (60.0) | 0.7 |
| Neoadjuvant Chemo Response (%) | 41 (54.7) | 8 (50.0) | 14 (53.8) | 19 (57.6) | 0.9 |
| Length of stay - days, mean (SD) | 8.10 (6.04) | 7.68 (4.76) | 8.21 (5.78) | 8.25 (6.92) | 0.9 |
| Length of Stay - days, median [IQR] | 7 [5.25, 8] | 7 [6, 7.5] | 6.5 [5, 8] | 7 [6, 8] | >0.9 |
| Operative time - hours, mean (SD) | 7.90 (2.26) | 8.17 (2.26) | 8.31 (2.18) | 7.27 (2.27) | 0.1 |
| Operative time - hours, median [IQR] | 8.12 [5.89, 9.63] | 8.17 [6.43, 9.95] | 8.37 [6.56, 9.67] | 6.90 [5.47, 9.04] | 0.1 |
| **Intravesical Treatment (%)** |  |  |  |  | 0.3 |
| None | 96 (71.6) | 22 (71.0) | 34 (70.8) | 40 (72.7) |  |
| BCG^c^ | 23 (17.2) | 8 (25.8) | 9 (18.8) | 6 (10.9) |  |
| Chemo | 8 (6.0) | 0 (0.0) | 2 (4.2) | 6 (10.9) |  |
| Both | 7 (5.2) | 1 (3.2) | 3 (6.2) | 3 (5.5) |  |
| **Systemic Treatment (%)** |  |  |  |  | 0.8 |
| None | 49 (36.6) | 13 (41.9) | 17 (35.4) | 19 (34.5) |  |
| Chemo | 73 (54.5) | 15 (48.4) | 26 (54.2) | 32 (58.2) |  |
| Immunotherapy | 1 (0.7) | 0 (0.0) | 0 (0.0) | 1 (1.8) |  |
| Both | 11 (8.2) | 3 (9.7) | 5 (10.4) | 3 (5.5) |  |
| **Complication (30 day)** |  |  |  |  |  |
| Readmission (%) | 50 (37.3) | 11 (35.5) | 19 (39.6) | 20 (36.4) | 0.9 |
| Ileus (%) | 23 (17.2) | 3 (9.7) | 9 (18.8) | 11 (20.0) | 0.4 |
| Post-operative Transfusion (%) | 28 (20.9) | 8 (25.8) | 13 (27.1) | 7 (12.7) | 0.2 |
| DVT/PE^d^ (%) | 12 (9.0) | 4 (12.9) | 3 (6.2) | 5 (9.1) | 0.6 |
| Reoperation (%) | 9 (6.7) | 2 (6.5) | 5 (10.4) | 2 (3.6) | 0.4 |
| **Pathology, T Stage (%)** |  |  |  |  | 0.6 |
| T0 | 27 (20.1) | 8 (25.8) | 6 (12.5) | 13 (23.6) |  |
| Ta | 8 (6.0) | 2 (6.5) | 3 (6.2) | 3 (5.5) |  |
| Tis | 8 (6.0) | 1 (3.2) | 2 (4.2) | 5 (9.1) |  |
| T1 | 15 (11.2) | 5 (16.1) | 3 (6.2) | 7 (12.7) |  |
| T2 | 31 (23.1) | 4 (12.9) | 15 (31.2) | 12 (21.8) |  |
| T3 | 33 (24.6) | 9 (29.0) | 14 (29.2) | 10 (18.2) |  |
| T4 | 12 (9.0) | 2 (6.5) | 5 (10.4) | 5 (9.1) |  |
| **Select Mutations (%)** |  |  |  |  |  |
| Matched Oncopanel (%) | 17 (12.7) | 6 (19.4) | 6 (12.5) | 5 (9.1) | 0.4 |
| TP53 | 11 (64.7) | 5 (83.3) | 5 (83.3) | 1 (20.0) | 0.045 |
| KMT2D | 6 (35.3) | 5 (83.3) | 1 (16.7) | 0 (0.0) | 0.008 |
| ARID1A | 4 (23.5) | 2 (33.3) | 1 (16.7) | 1 (20.0) | 0.8 |
| RB1 | 4 (23.5) | 1 (16.7) | 2 (33.3) | 1 (20.0) | 0.8 |
| TERT | 4 (23.5) | 1 (16.7) | 2 (33.3) | 1 (20.0) | 0.8 |
| KDM6A | 4 (23.5) | 0 (0.0) | 2 (33.3) | 2 (40.0) | 0.2 |
| PIK3CA | 3 (17.6) | 3 (50.0) | 0 (0.0) | 0 (0.0) | 0.035 |
| CDKN1A | 2 (11.8) | 1 (16.7) | 0 (0.0) | 1 (20.0) | 0.5 |
| FGFR3 | 1 (5.9) | 0 (0.0) | 0 (0.0) | 1 (20.0) | 0.3 |
| CDKN2A | 1 (5.9) | 0 (0.0) | 1 (16.7) | 0 (0.0) | 0.4 |
| STAG2 | 1 (5.9) | 0 (0.0) | 1 (16.7) | 0 (0.0) | 0.4 |
| CDKN2B | 0 (0.0) | 0 (0.0) | 0 (0.0) | 0 (0.0) | NA |
| CCND1 | 0 (0.0) | 0 (0.0) | 0 (0.0) | 0 (0.0) | NA |
| ERBB2 | 0 (0.0) | 0 (0.0) | 0 (0.0) | 0 (0.0) | NA |
| EP300 | 0 (0.0) | 0 (0.0) | 0 (0.0) | 0 (0.0) | NA |

*a = Mass General Brigham Young Cystectomy Cohort (MGB-YCC). b = Body Mass Index (BMI). c = Bacillus Calmette-Guérin (BCG) Therapy. d = Deep Vein Thrombosis/Pulmonary Embolism (DVT/PE).*
