## Supplementary material for "Clinical and genomic profiling of early-onset bladder cancer identifies key alterations and therapeutic targets": Table S2

**Table S2: Detailed MSK-IMPACT^a^ Patient Characteristics**

| **Age** | **Overall** | **< 55** | **≥ 55** | **p** | **< 45** | **45 - 49** | **50 - 54** | **p** |
| --- | --- | --- | --- | --- | --- | --- | --- | --- |
| N (%) | 1271 (100.0) | 212 (16.7) | 1059 (83.3) |  | 60 (28.3) | 56 (26.4) | 96 (45.3) |  |
| Age, mean (SD) | 65.22 (11.12) | 47.16 (6.94) | 68.84 (7.77) | <0.001 | 37.75 (5.22) | 47.77 (1.48) | 52.67 (1.38) | <0.001 |
| Age, median [IQR] | 66.4 [58.4, 73.2] | 49.2 [43.4, 52.3] | 68.40 [62.40, 74.60] | <0.001 | 38.2 [34.6, 42.5] | 48.1 [46.7, 49.0] | 52.8 [51.9, 53.7] | <0.001 |
| Male (%) | 980 (77.1) | 160 (75.5) | 820 (77.4) | 0.6 | 45 (75.0) | 48 (85.7) | 67 (69.8) | 0.08 |
| Smoking (%) | 813 (64.7) | 111 (53.1) | 702 (67.0) | <0.001 | 22 (36.7) | 28 (51.9) | 61 (64.2) | 0.004 |
| Stage ≥T2, Muscle Invasive (%) | 777 (61.1) | 115 (54.2) | 662 (62.5) | 0.03 | 28 (46.7) | 27 (48.2) | 60 (62.5) | 0.09 |
| **Intravesical Treatment (%)** |  |  |  | 0.3 |  |  |  | 0.8 |
| None | 891 (70.2) | 142 (67.3) | 749 (70.8) |  | 44 (73.3) | 37 (66.1) | 61 (64.2) |  |
| BCG^b^ | 257 (20.3) | 46 (21.8) | 211 (19.9) |  | 11 (18.3) | 14 (25.0) | 21 (22.1) |  |
| Chemo | 30 (2.4) | 3 (1.4) | 27 (2.6) |  | 1 (1.7) | 0 (0.0) | 2 (2.1) |  |
| Both | 91 (7.2) | 20 (9.5) | 71 (6.7) |  | 4 (6.7) | 5 (8.9) | 11 (11.6) |  |
| **Systemic Treatment (%)** |  |  |  | 0.2 |  |  |  | NA |
| None | 996 (78.9) | 164 (78.5) | 832 (79.0) |  | 50 (83.3) | 39 (72.2) | 75 (78.9) |  |
| Chemo | 220 (17.4) | 42 (20.1) | 178 (16.9) |  | 9 (15.0) | 13 (24.1) | 20 (21.1) |  |
| Immunotherapy | 16 (1.3) | 0 (0.0) | 16 (1.5) |  | 0 (0.0) | 0 (0.0) | 0 (0.0) |  |
| Both | 30 (2.4) | 3 (1.4) | 27 (2.6) |  | 1 (1.7) | 2 (3.7) | 0 (0.0) |  |
| **Specimen Stage (%)** |  |  |  | 0.001 |  |  |  | 0.01 |
| Low Grade | 69 (5.4) | 22 (10.4) | 47 (4.4) |  | 12 (20.0) | 5 (8.9) | 5 (5.2) |  |
| High Grade, Non-Invasive | 234 (18.4) | 48 (22.6) | 186 (17.6) |  | 11 (18.3) | 18 (32.1) | 19 (19.8) |  |
| High Grade, Invasive | 737 (58.0) | 105 (49.5) | 632 (59.7) |  | 32 (53.3) | 22 (39.3) | 51 (53.1) |  |
| Metastatic | 231 (18.2) | 37 (17.5) | 194 (18.3) |  | 5 (8.3) | 11 (19.6) | 21 (21.9) |  |
| **Specimen Type (%)** |  |  |  | 0.08 |  |  |  | NA |
| Cystectomy | 207 (16.3) | 27 (12.7) | 180 (17.0) |  | 3 (5.0) | 8 (14.3) | 16 (16.7) |  |
| Metastasis | 231 (18.2) | 37 (17.5) | 194 (18.3) |  | 5 (8.3) | 11 (19.6) | 21 (21.9) |  |
| Partial Cystectomy | 14 (1.1) | 2 (0.9) | 12 (1.1) |  | 0 (0.0) | 1 (1.8) | 1 (1.0) |  |
| TUR | 811 (63.8) | 143 (67.5) | 668 (63.1) |  | 51 (85.0) | 36 (64.3) | 56 (58.3) |  |
| Urethral biopsy | 3 (0.2) | 0 (0.0) | 3 (0.3) |  | 0 (0.0) | 0 (0.0) | 0 (0.0) |  |
| Urethrectomy | 5 (0.4) | 3 (1.4) | 2 (0.2) |  | 1 (1.7) | 0 (0.0) | 2 (2.1) |  |
| **Stage (%)** |  |  |  | 0.006 |  |  |  | 0.006 |
| Low Grade | 69 (5.4) | 22 (10.4) | 47 (4.4) |  | 12 (20.0) | 5 (8.9) | 5 (5.2) |  |
| High Grade | 378 (29.7) | 65 (30.7) | 313 (29.6) |  | 17 (28.3) | 24 (42.9) | 24 (25.0) |  |
| CIS^c^ | 47 (3.7) | 10 (4.7) | 37 (3.5) |  | 3 (5.0) | 0 (0.0) | 7 (7.3) |  |
| Muscle-Invasive | 546 (43.0) | 78 (36.8) | 468 (44.2) |  | 23 (38.3) | 16 (28.6) | 39 (40.6) |  |
| Metastatic | 231 (18.2) | 37 (17.5) | 194 (18.3) |  | 5 (8.3) | 11 (19.6) | 21 (21.9) |  |
| **Histology (%)** |  |  |  | 0.3 |  |  |  | 0.4 |
| Urothelial Carcinoma | 936 (73.6) | 167 (78.8) | 769 (72.6) |  | 50 (83.3) | 49 (87.5) | 68 (70.8) |  |
| Squamous Cell | 84 (6.6) | 16 (7.5) | 68 (6.4) |  | 5 (8.3) | 2 (3.6) | 9 (9.4) |  |
| Plasmacytoid | 37 (2.9) | 5 (2.4) | 32 (3.0) |  | 0 (0.0) | 1 (1.8) | 4 (4.2) |  |
| Small Cell, Neuroendocrine | 49 (3.9) | 7 (3.3) | 42 (4.0) |  | 1 (1.7) | 1 (1.8) | 5 (5.2) |  |
| Micropapillary | 40 (3.1) | 2 (0.9) | 38 (3.6) |  | 0 (0.0) | 0 (0.0) | 2 (2.1) |  |
| Glandular | 20 (1.6) | 2 (0.9) | 18 (1.7) |  | 0 (0.0) | 0 (0.0) | 2 (2.1) |  |
| Other | 105 (8.3) | 13 (6.1) | 92 (8.7) |  | 4 (6.7) | 3 (5.4) | 6 (6.2) |  |
| **Select Mutations (%)** |  |  |  |  |  |  |  |  |
| TERT | 937 (73.7) | 144 (67.9) | 793 (74.9) | 0.044 | 34 (56.7) | 34 (60.7) | 76 (79.2) | 0.006 |
| TP53 | 612 (48.2) | 85 (40.1) | 527 (49.8) | 0.013 | 21 (35.0) | 17 (30.4) | 47 (49.0) | 0.05 |
| FGFR3 | 319 (25.1) | 78 (36.8) | 241 (22.8) | <0.001 | 26 (43.3) | 21 (37.5) | 31 (32.3) | 0.4 |
| KDM6A | 332 (26.1) | 52 (24.5) | 280 (26.4) | 0.6 | 14 (23.3) | 13 (23.2) | 25 (26.0) | 0.9 |
| PIK3CA | 236 (18.6) | 46 (21.7) | 190 (17.9) | 0.2 | 12 (20.0) | 9 (16.1) | 25 (26.0) | 0.3 |
| CDKN2A | 285 (22.4) | 41 (19.3) | 244 (23.0) | 0.3 | 9 (15.0) | 12 (21.4) | 20 (20.8) | 0.6 |
| ARID1A | 299 (23.5) | 39 (18.4) | 260 (24.6) | 0.07 | 13 (21.7) | 7 (12.5) | 19 (19.8) | 0.4 |
| CDKN2B | 218 (17.2) | 36 (17.0) | 182 (17.2) | 0.9 | 9 (15.0) | 9 (16.1) | 18 (18.8) | 0.8 |
| KMT2D | 235 (18.5) | 31 (14.6) | 204 (19.3) | 0.1 | 10 (16.7) | 5 (8.9) | 16 (16.7) | 0.4 |
| CDKN1A | 150 (11.8) | 27 (12.7) | 123 (11.6) | 0.7 | 11 (18.3) | 6 (10.7) | 10 (10.4) | 0.3 |
| RB1 | 243 (19.1) | 26 (12.3) | 217 (20.5) | 0.007 | 6 (10.0) | 2 (3.6) | 18 (18.8) | 0.019 |
| CCND1 | 139 (10.9) | 26 (12.3) | 113 (10.7) | 0.6 | 7 (11.7) | 9 (16.1) | 10 (10.4) | 0.6 |
| ERBB2 | 192 (15.1) | 24 (11.3) | 168 (15.9) | 0.1 | 5 (8.3) | 3 (5.4) | 16 (16.7) | 0.07 |
| STAG2 | 123 (9.7) | 23 (10.8) | 100 (9.4) | 0.6 | 9 (15.0) | 4 (7.1) | 10 (10.4) | 0.4 |
| EP300 | 92 (7.2) | 19 (9.0) | 73 (6.9) | 0.4 | 4 (6.7) | 3 (5.4) | 12 (12.5) | 0.3 |

*a = Memorial Sloan Kettering Integrated Mutation Profiling of Actionable Cancer Targets (MSK-IMPACT) Cohort. b = Bacillus Calmette-Guérin (BCG) Therapy. c = carcinoma in situ (CIS).*
