## Supplementary material for "Clinical and genomic profiling of early-onset bladder cancer identifies key alterations and therapeutic targets": Table S3

**Table S3: Variant Classifications for Most Frequent Mutations by Cohort**

**A) MGB-YCC^a^**

| **Hugo Symbol** | **Frame Shift Deletion** | **Frame Shift Insertion** | **In Frame Insertion** | **Missense** | **Nonsense** | **Splice Site** | **Total** | **Mutated Samples** | **Altered Samples** |
| --- | --- | --- | --- | --- | --- | --- | --- | --- | --- |
| TP53 | 2 | 0 | 0 | 8 | 3 | 1 | 14 | 11 | 11 |
| KMT2D | 2 | 0 | 0 | 11 | 1 | 1 | 15 | 6 | 6 |
| KDM6A | 0 | 1 | 0 | 1 | 1 | 1 | 4 | 4 | 4 |
| RB1 | 1 | 0 | 0 | 0 | 2 | 1 | 4 | 4 | 4 |
| NOTCH2 | 0 | 0 | 0 | 4 | 0 | 1 | 5 | 3 | 3 |
| ARID2 | 0 | 0 | 0 | 4 | 0 | 0 | 4 | 3 | 3 |
| ATRX | 0 | 0 | 0 | 1 | 2 | 1 | 4 | 3 | 3 |
| SLX4 | 0 | 0 | 0 | 4 | 0 | 0 | 4 | 3 | 3 |
| ARID1A | 1 | 0 | 0 | 0 | 2 | 0 | 3 | 3 | 3 |
| ATM | 0 | 0 | 0 | 2 | 1 | 0 | 3 | 3 | 3 |
| BRCA1 | 0 | 0 | 0 | 3 | 0 | 0 | 3 | 3 | 3 |
| CHEK2 | 2 | 0 | 0 | 0 | 1 | 0 | 3 | 3 | 3 |
| CIITA | 0 | 0 | 0 | 3 | 0 | 0 | 3 | 3 | 3 |
| CREBBP | 0 | 0 | 0 | 2 | 1 | 0 | 3 | 3 | 3 |
| CUX1 | 0 | 0 | 0 | 3 | 0 | 0 | 3 | 3 | 3 |
| NOTCH1 | 0 | 0 | 0 | 3 | 0 | 0 | 3 | 3 | 3 |
| PIK3CA | 0 | 0 | 0 | 3 | 0 | 0 | 3 | 3 | 3 |
| CDKN1A | 0 | 1 | 0 | 0 | 2 | 0 | 3 | 2 | 2 |
| ERCC6 | 0 | 0 | 1 | 2 | 0 | 0 | 3 | 2 | 2 |

**B) MSK-IMPACT^b^ Cystectomy Samples (all ages)**

| **Hugo Symbol** | **Frame Shift Deletion** | **Frame Shift Insertion** | **In Frame Deletion** | **Missense** | **Nonsense** | **Splice Site** | **Translation Start Site** | **Total** | **Mutated Samples** | **Altered Samples** |
| --- | --- | --- | --- | --- | --- | --- | --- | --- | --- | --- |
| TP53 | 1 | 0 | 3 | 55 | 7 | 2 | 0 | 68 | 63 | 63 |
| FGFR3 | 0 | 0 | 0 | 30 | 0 | 0 | 0 | 30 | 29 | 29 |
| PIK3CA | 0 | 0 | 0 | 26 | 0 | 0 | 0 | 26 | 24 | 24 |
| CDKN2A | 1 | 0 | 0 | 9 | 2 | 0 | 0 | 12 | 12 | 12 |
| ERBB2 | 0 | 0 | 0 | 12 | 0 | 0 | 0 | 12 | 12 | 12 |
| RXRA | 0 | 0 | 0 | 9 | 0 | 0 | 0 | 9 | 9 | 9 |
| ATM | 0 | 0 | 0 | 11 | 1 | 0 | 0 | 12 | 8 | 8 |
| ERBB3 | 0 | 0 | 0 | 7 | 0 | 0 | 0 | 7 | 7 | 7 |
| NOTCH3 | 0 | 1 | 0 | 6 | 1 | 0 | 0 | 8 | 5 | 5 |
| KMT2D | 1 | 0 | 0 | 1 | 4 | 0 | 0 | 6 | 5 | 5 |
| RB1 | 0 | 0 | 0 | 4 | 1 | 0 | 0 | 5 | 5 | 5 |
| FAT1 | 0 | 0 | 0 | 5 | 0 | 0 | 0 | 5 | 4 | 4 |
| FOXP1 | 0 | 1 | 0 | 1 | 2 | 0 | 0 | 4 | 4 | 4 |
| KRAS | 0 | 0 | 0 | 4 | 0 | 0 | 0 | 4 | 4 | 4 |
| PTPRT | 0 | 0 | 0 | 4 | 0 | 0 | 0 | 4 | 4 | 4 |
| RHOA | 0 | 0 | 0 | 4 | 0 | 0 | 0 | 4 | 4 | 4 |
| ASXL2 | 0 | 0 | 0 | 3 | 0 | 0 | 0 | 3 | 3 | 3 |
| CREBBP | 1 | 0 | 0 | 2 | 0 | 0 | 0 | 3 | 3 | 3 |
| DOT1L | 0 | 0 | 0 | 3 | 0 | 0 | 0 | 3 | 3 | 3 |

**C) MSK-IMPACT^b^ Cystectomy Samples (early-onset cases)**

| **Hugo Symbol** | **Frame Shift Deletion** | **Frame Shift Insertion** | **Missense** | **Nonsense** | **Splice Site** | **Translation Start Site** | **Total** | **Mutated Samples** | **Altered Samples** |
| --- | --- | --- | --- | --- | --- | --- | --- | --- | --- |
| FGFR3 | 0 | 0 | 8 | 0 | 0 | 0 | 8 | 8 | 8 |
| TP53 | 0 | 0 | 4 | 1 | 1 | 0 | 6 | 6 | 6 |
| PIK3CA | 0 | 0 | 6 | 0 | 0 | 0 | 6 | 5 | 5 |
| ATM | 0 | 0 | 5 | 0 | 0 | 0 | 5 | 2 | 2 |
| B2M | 0 | 0 | 1 | 0 | 0 | 1 | 2 | 2 | 2 |
| ERBB2 | 0 | 0 | 2 | 0 | 0 | 0 | 2 | 2 | 2 |
| ERBB3 | 0 | 0 | 2 | 0 | 0 | 0 | 2 | 2 | 2 |
| JAK3 | 1 | 0 | 1 | 0 | 0 | 0 | 2 | 2 | 2 |
| MEF2B | 0 | 0 | 2 | 0 | 0 | 0 | 2 | 2 | 2 |
| RB1 | 0 | 0 | 2 | 0 | 0 | 0 | 2 | 2 | 2 |
| TET2 | 0 | 0 | 2 | 0 | 0 | 0 | 2 | 2 | 2 |
| NOTCH3 | 0 | 0 | 2 | 1 | 0 | 0 | 3 | 1 | 1 |
| APC | 0 | 0 | 2 | 0 | 0 | 0 | 2 | 1 | 1 |
| JAK1 | 0 | 0 | 2 | 0 | 0 | 0 | 2 | 1 | 1 |
| KMT2D | 0 | 0 | 0 | 2 | 0 | 0 | 2 | 1 | 1 |
| ARAF | 0 | 0 | 1 | 0 | 0 | 0 | 1 | 1 | 1 |
| BAP1 | 0 | 0 | 1 | 0 | 0 | 0 | 1 | 1 | 1 |
| BCL6 | 0 | 0 | 1 | 0 | 0 | 0 | 1 | 1 | 1 |
| BRIP1 | 0 | 0 | 1 | 0 | 0 | 0 | 1 | 1 | 1 |

**D) MSK-IMPACT^b^ Cystectomy Samples (late-onset cases)**

| **Hugo Symbol** | **Frame Shift Deletion** | **Frame Shift Insertion** | **In Frame Deletion** | **Missense** | **Nonsense** | **Splice Site** | **Total** | **Mutated Samples** | **Altered Samples** |
| --- | --- | --- | --- | --- | --- | --- | --- | --- | --- |
| TP53 | 1 | 0 | 3 | 51 | 6 | 1 | 62 | 57 | 57 |
| FGFR3 | 0 | 0 | 0 | 22 | 0 | 0 | 22 | 21 | 21 |
| PIK3CA | 0 | 0 | 0 | 20 | 0 | 0 | 20 | 19 | 19 |
| CDKN2A | 1 | 0 | 0 | 9 | 2 | 0 | 12 | 12 | 12 |
| ERBB2 | 0 | 0 | 0 | 10 | 0 | 0 | 10 | 10 | 10 |
| RXRA | 0 | 0 | 0 | 8 | 0 | 0 | 8 | 8 | 8 |
| ATM | 0 | 0 | 0 | 6 | 1 | 0 | 7 | 6 | 6 |
| ERBB3 | 0 | 0 | 0 | 5 | 0 | 0 | 5 | 5 | 5 |
| NOTCH3 | 0 | 1 | 0 | 4 | 0 | 0 | 5 | 4 | 4 |
| KMT2D | 1 | 0 | 0 | 1 | 2 | 0 | 4 | 4 | 4 |
| FAT1 | 0 | 0 | 0 | 4 | 0 | 0 | 4 | 3 | 3 |
| ASXL2 | 0 | 0 | 0 | 3 | 0 | 0 | 3 | 3 | 3 |
| FOXP1 | 0 | 1 | 0 | 0 | 2 | 0 | 3 | 3 | 3 |
| KRAS | 0 | 0 | 0 | 3 | 0 | 0 | 3 | 3 | 3 |
| MSH6 | 0 | 0 | 0 | 3 | 0 | 0 | 3 | 3 | 3 |
| NOTCH4 | 1 | 0 | 0 | 2 | 0 | 0 | 3 | 3 | 3 |
| PTPRT | 0 | 0 | 0 | 3 | 0 | 0 | 3 | 3 | 3 |
| RB1 | 0 | 0 | 0 | 2 | 1 | 0 | 3 | 3 | 3 |
| RHOA | 0 | 0 | 0 | 3 | 0 | 0 | 3 | 3 | 3 |

**E) MSK-IMPACT^b^ Transurethral Resection Samples (all ages)**

| **Hugo Symbol** | **Frame Shift Deletion** | **Frame Shift Insertion** | **In Frame Deletion** | **In Frame Insertion** | **Missense** | **Nonsense** | **Splice Site** | **Total** | **Mutated Samples** | **Altered Samples** |
| --- | --- | --- | --- | --- | --- | --- | --- | --- | --- | --- |
| TP53 | 4 | 1 | 0 | 0 | 225 | 30 | 9 | 269 | 229 | 229 |
| FGFR3 | 0 | 0 | 0 | 0 | 234 | 0 | 0 | 234 | 224 | 224 |
| PIK3CA | 0 | 0 | 0 | 0 | 163 | 0 | 0 | 163 | 150 | 150 |
| ERBB2 | 0 | 0 | 0 | 0 | 66 | 0 | 0 | 66 | 59 | 59 |
| KRAS | 0 | 0 | 0 | 0 | 32 | 0 | 0 | 32 | 31 | 31 |
| RXRA | 0 | 1 | 0 | 0 | 25 | 0 | 0 | 26 | 26 | 26 |
| ERBB3 | 0 | 0 | 0 | 0 | 20 | 0 | 0 | 20 | 18 | 18 |
| ATM | 0 | 0 | 0 | 0 | 15 | 2 | 1 | 18 | 17 | 17 |
| KMT2D | 1 | 0 | 0 | 0 | 10 | 8 | 0 | 19 | 16 | 16 |
| RB1 | 0 | 0 | 0 | 0 | 3 | 13 | 2 | 18 | 16 | 16 |
| CDKN2A | 0 | 0 | 0 | 0 | 12 | 5 | 0 | 17 | 16 | 16 |
| RHOA | 0 | 0 | 0 | 0 | 15 | 0 | 0 | 15 | 15 | 15 |
| CTNNB1 | 0 | 0 | 0 | 0 | 14 | 0 | 0 | 14 | 14 | 14 |
| FBXW7 | 0 | 0 | 0 | 0 | 13 | 0 | 0 | 13 | 13 | 13 |
| HRAS | 0 | 0 | 0 | 0 | 12 | 0 | 0 | 12 | 12 | 12 |
| CREBBP | 0 | 0 | 1 | 0 | 8 | 3 | 0 | 12 | 11 | 11 |
| AKT1 | 0 | 0 | 0 | 0 | 11 | 0 | 0 | 11 | 11 | 11 |
| ARID1A | 1 | 1 | 1 | 0 | 4 | 4 | 0 | 11 | 11 | 11 |
| BRAF | 0 | 0 | 0 | 0 | 11 | 0 | 0 | 11 | 11 | 11 |

**F) MSK-IMPACT^b^ Transurethral Resection Samples (early-onset cases)**

| **Hugo Symbol** | **Frame Shift Deletion** | **Frame Shift Insertion** | **In Frame Insertion** | **Missense** | **Nonsense** | **Splice Site** | **Total** | **Mutated Samples** | **Altered Samples** |
| --- | --- | --- | --- | --- | --- | --- | --- | --- | --- |
| FGFR3 | 0 | 0 | 0 | 56 | 0 | 0 | 56 | 53 | 53 |
| TP53 | 0 | 0 | 0 | 31 | 8 | 2 | 41 | 39 | 39 |
| PIK3CA | 0 | 0 | 0 | 30 | 0 | 0 | 30 | 27 | 27 |
| U2AF1 | 0 | 0 | 0 | 5 | 0 | 0 | 5 | 5 | 5 |
| ERBB2 | 0 | 0 | 0 | 5 | 0 | 0 | 5 | 4 | 4 |
| KRAS | 0 | 0 | 0 | 5 | 0 | 0 | 5 | 4 | 4 |
| FBXW7 | 0 | 0 | 0 | 4 | 0 | 0 | 4 | 4 | 4 |
| RNF43 | 2 | 0 | 0 | 2 | 0 | 0 | 4 | 4 | 4 |
| ARID1A | 0 | 0 | 0 | 1 | 2 | 0 | 3 | 3 | 3 |
| BRAF | 0 | 0 | 0 | 3 | 0 | 0 | 3 | 3 | 3 |
| CREBBP | 0 | 0 | 0 | 3 | 0 | 0 | 3 | 3 | 3 |
| CTCF | 0 | 3 | 0 | 0 | 0 | 0 | 3 | 3 | 3 |
| CTNNB1 | 0 | 0 | 0 | 3 | 0 | 0 | 3 | 3 | 3 |
| ERBB3 | 0 | 0 | 0 | 3 | 0 | 0 | 3 | 3 | 3 |
| KDM5A | 2 | 0 | 0 | 1 | 0 | 0 | 3 | 3 | 3 |
| PTPRT | 0 | 0 | 0 | 2 | 1 | 0 | 3 | 3 | 3 |
| RIT1 | 0 | 0 | 0 | 3 | 0 | 0 | 3 | 3 | 3 |
| RXRA | 0 | 0 | 0 | 3 | 0 | 0 | 3 | 3 | 3 |
| TET2 | 0 | 0 | 0 | 3 | 0 | 0 | 3 | 3 | 3 |

**G) MSK-IMPACT^b^ Transurethral Resection Samples (late-onset cases)**

| **Hugo Symbol** | **Frame Shift Deletion** | **Frame Shift Insertion** | **In Frame Deletion** | **In Frame Insertion** | **Missense** | **Nonsense** | **Splice Site** | **Total** | **Mutated Samples** | **Altered Samples** |
| --- | --- | --- | --- | --- | --- | --- | --- | --- | --- | --- |
| TP53 | 4 | 1 | 0 | 0 | 194 | 22 | 7 | 228 | 190 | 190 |
| FGFR3 | 0 | 0 | 0 | 0 | 178 | 0 | 0 | 178 | 171 | 171 |
| PIK3CA | 0 | 0 | 0 | 0 | 133 | 0 | 0 | 133 | 123 | 123 |
| ERBB2 | 0 | 0 | 0 | 0 | 61 | 0 | 0 | 61 | 55 | 55 |
| KRAS | 0 | 0 | 0 | 0 | 27 | 0 | 0 | 27 | 27 | 27 |
| RXRA | 0 | 1 | 0 | 0 | 22 | 0 | 0 | 23 | 23 | 23 |
| KMT2D | 1 | 0 | 0 | 0 | 10 | 8 | 0 | 19 | 16 | 16 |
| ATM | 0 | 0 | 0 | 0 | 14 | 2 | 1 | 17 | 16 | 16 |
| ERBB3 | 0 | 0 | 0 | 0 | 17 | 0 | 0 | 17 | 15 | 15 |
| RB1 | 0 | 0 | 0 | 0 | 3 | 12 | 2 | 17 | 15 | 15 |
| CDKN2A | 0 | 0 | 0 | 0 | 11 | 5 | 0 | 16 | 15 | 15 |
| RHOA | 0 | 0 | 0 | 0 | 13 | 0 | 0 | 13 | 13 | 13 |
| AKT1 | 0 | 0 | 0 | 0 | 11 | 0 | 0 | 11 | 11 | 11 |
| CTNNB1 | 0 | 0 | 0 | 0 | 11 | 0 | 0 | 11 | 11 | 11 |
| HRAS | 0 | 0 | 0 | 0 | 11 | 0 | 0 | 11 | 11 | 11 |
| FBXW7 | 0 | 0 | 0 | 0 | 9 | 0 | 0 | 9 | 9 | 9 |
| CREBBP | 0 | 0 | 1 | 0 | 5 | 3 | 0 | 9 | 8 | 8 |
| ARID1A | 1 | 1 | 1 | 0 | 3 | 2 | 0 | 8 | 8 | 8 |
| BRAF | 0 | 0 | 0 | 0 | 8 | 0 | 0 | 8 | 8 | 8 |

*a = Mass General Brigham Young Cystectomy Cohort (MGB-YCC). b = Memorial Sloan Kettering Integrated Mutation Profiling of Actionable Cancer Targets (MSK-IMPACT) Cohort.*
