## Supplementary figures and images for "Clinical and genomic profiling of early-onset bladder cancer identifies key alterations and therapeutic targets"

### Figure S1

#### Mass General Brigham Young Cystectomy Cohort

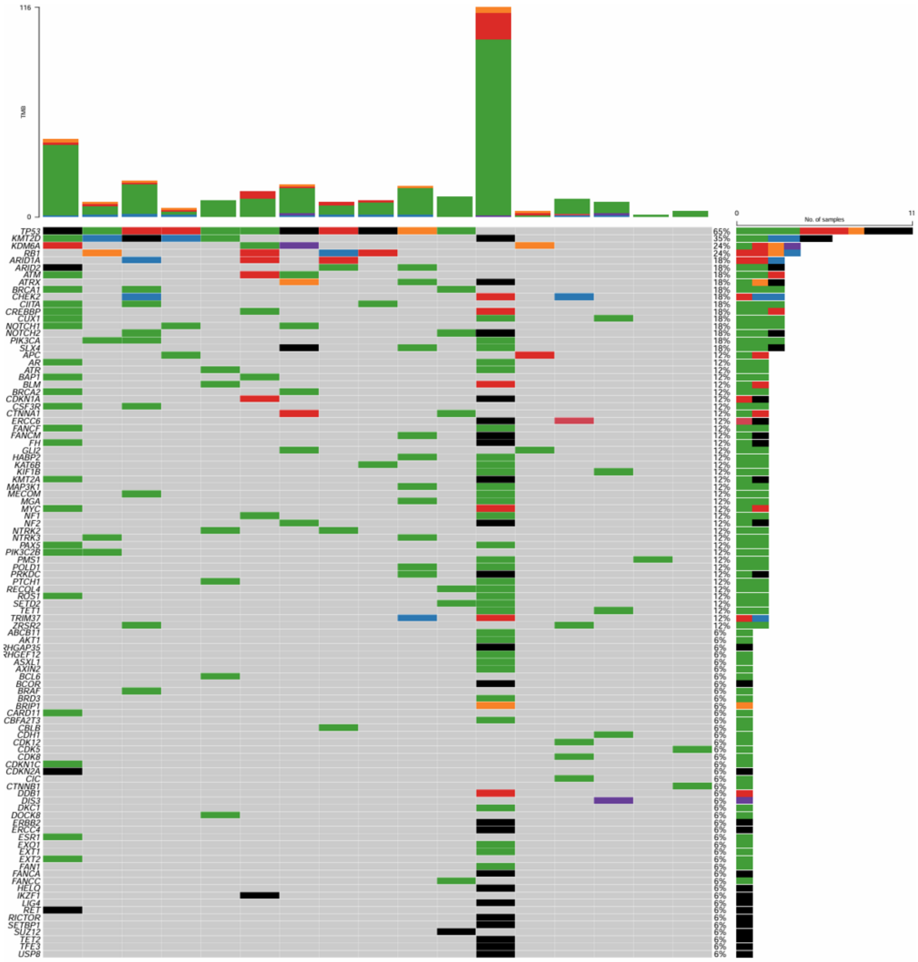


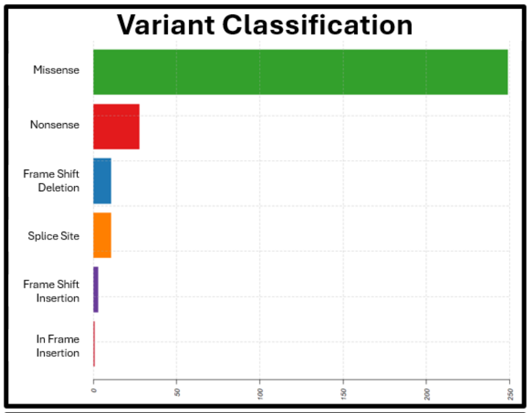


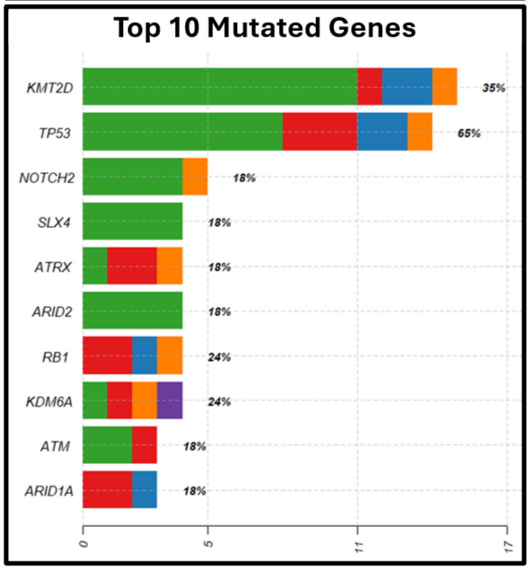
