## Supplementary material for "Clinical and genomic profiling of early-onset bladder cancer identifies key alterations and therapeutic targets": Figure S2

### MSK-IMPACT: Cystectomy Samples from Early-Onset Patients

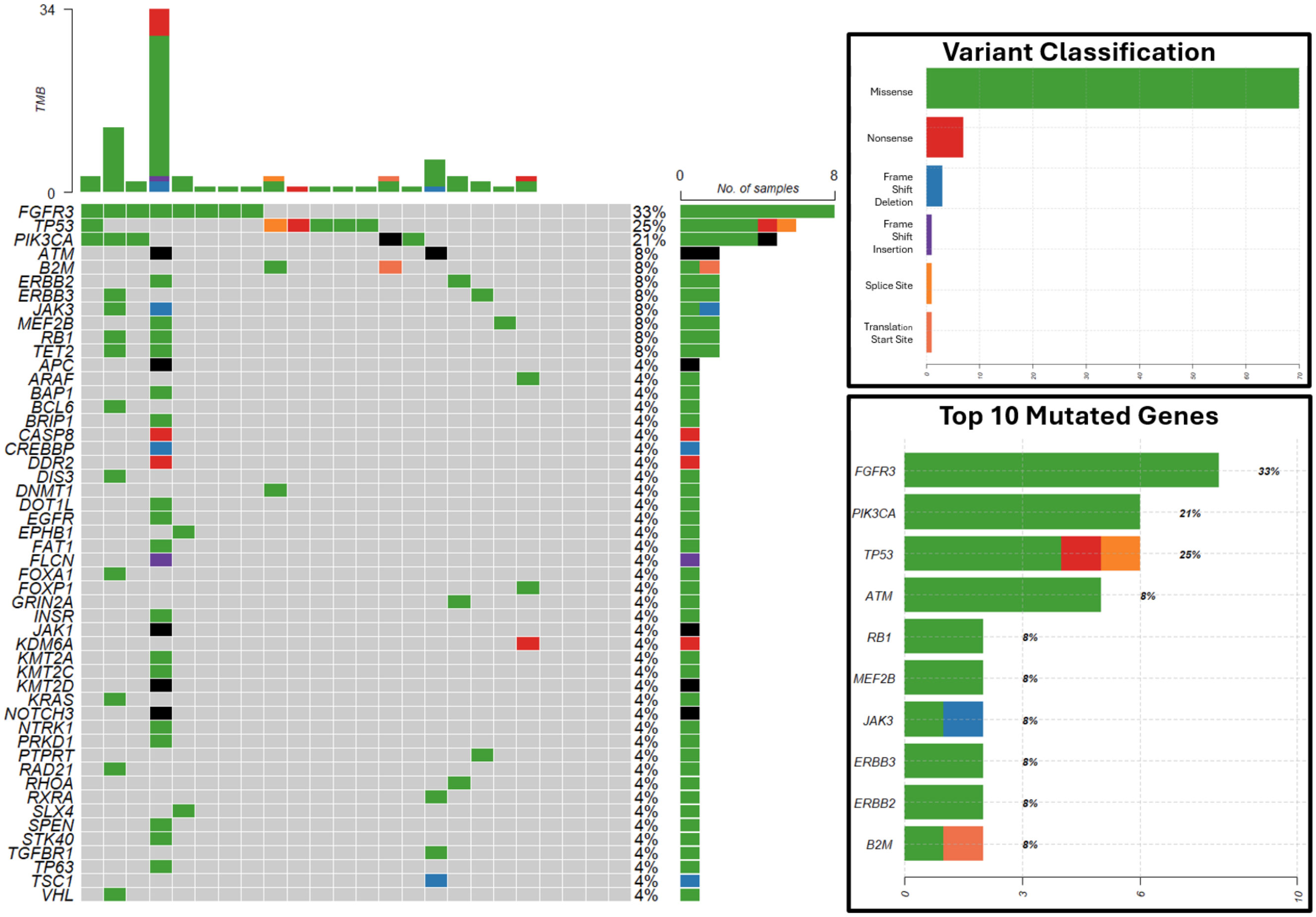


### MSK-IMPACT: Cystectomy Samples from Late-Onset Patients

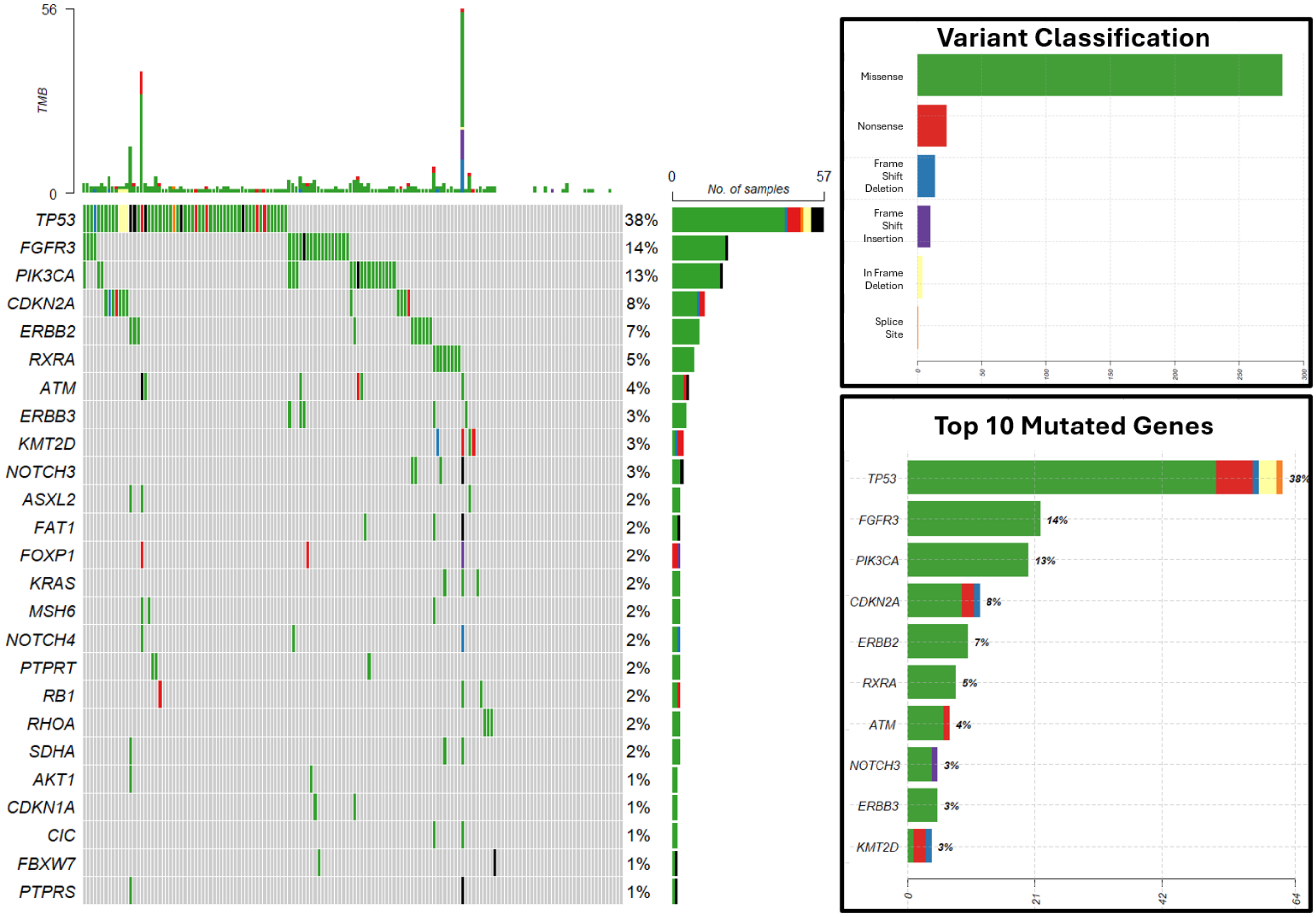
